# Temporal dynamics of asymptomatic malaria in a high transmission community in Ghana

**DOI:** 10.64898/2026.07.11.26357819

**Authors:** Cecil Kwame Mfum Asa-Atiemo, Dorothy Agbo, Festus Kojo Acquah, Evans Kofi Obboh, Anne Poinsignon, Rachida Tahar, Linda Eva Amoah

**Author notes:** Corresponding Author: Linda Eva Amoah^1,5 (^^)^.

## Abstract

Asymptomatic *Plasmodium falciparum* carriers are cryptic reservoirs that sustain malaria transmission, undermining control efforts. However, its temporal dynamics and drivers remain underexplored. A total of 845 asymptomatic individuals in Simiw were screened for *P. falciparum* carriage in May, August, and October 2021, as well as February 2022, representing the pre-peak, peak and post peak malaria transmission seasons that fall in either the wet or dry seasons. Meteorological data from March 2021 to March 2022 was obtained. Descriptive and inferential statistical analyses, such as Poisson Generalised Linear and Distributed Lag Nonlinear Models, were conducted in R to investigate the influence of seasonality and meteorological factors on PET-PCR-detected asymptomatic *Plasmodium falciparum* infections. In Simiw, the prevalence of asymptomatic *P. falciparum* infections ranged from 6.2% to 14.0% by RDT and 13.4% to 32.0% by PET-PCR during the study period. Asymptomatic malaria cases were highest in February 2022, during the dry season. Younger participants (median age: 14 years) had more infections than adults. Seasonal trend showed the strongest nonlinear relationship (exp(β) = 190.38, p = 0.01). Features of the dry season predicted asymptomatic malaria cases, as average maximum temperature (exp(β) = 1.30, p < 0.001) was a positive nonlinear predictor, whereas average humidity (exp(β) = 0.73, p = 0.02) and precipitation (exp(β) = 0.99, p < 0.001) were negative nonlinear predictors. According to regression models, the highest counts of asymptomatic malaria cases occur at a monthly maximum temperature of 32°C, monthly average relative humidity of 91% and monthly precipitation of 20 mm, similar to records in February 2022 in the dry season. Additionally, a precipitation of 150 mm forecasted the highest number of cases two months later (exp(β) = 3.20, p < 0.001). In this study, the seasonal trend had a much stronger effect on asymptomatic malaria than meteorological factors, such as rainfall.

## Introduction

Malaria is a deadly infectious disease caused by the parasite, *Plasmodium spp.* and transmitted by the vector, *Anopheles spp.* In Ghana, the most common etiologic species of malaria is *Plasmodium falciparum*, predominantly transmitted by *Anopheles gambiae sensu lato* and *Anopheles funestus* (1–3). *P. falciparum* may cause severe, mild, or asymptomatic malaria. Malaria imposes a substantial economic burden across Africa, costing 1.3% of the Gross Domestic Product (GDP) (4). Among at-risk populations in Ghana, malaria is detected in 35% of paediatric hospital admissions (5) and 11-26% of pregnant women (6,7). Reducing the malaria burden by 90% by 2030 would increase GDP by $16 billion per year (8).

The major temporal characteristic of malaria transmission is its seasonality, which is mainly determined by climatic or meteorological factors. This is because malaria transmission seasons follow rainfall patterns that give rise to the mosquito vector. Essentially, wet or dry seasons give rise to peak and off-peak malaria transmission seasons. There are two rainy seasons (bimodal rainfall) in southern Ghana, from April to June (first rainy season) and October to November (second rainy season), and one dry season from December to March every year, with atmospheric temperatures ranging from 23°C to 34°C, humidity ranging from 95% to 100%, and an average annual rainfall ranging from 787 mm to 830 mm. July and August are known as the little dry season (9,10). Following rainfall in southern Ghana, malaria incidence typically lags by one (1) month, peaking in June and July and lasting until November (2,5). Consistent with annual transmission patterns, there were 195 malaria cases per month in the wet season versus 133 cases per month in the dry season, occurring in 26.5% of children under 5 years, 58.7% of pregnant women, and 14.1% of asymptomatic people visiting hospitals in Ghana (11).

On the other hand, northern Ghana has one long rainy season from April to October every year (unimodal) (12,13). During the dry season, a dry northeastern wind of 38°C and 25% humidity blows, typically in January and February, compared to the average annual atmospheric temperature of 31°C (13). As a result of climate change in the country, the annual atmospheric temperature has increased by 1°C, and rainfall has decreased by 10% to 20% in the past 30 years. Additionally, there is an increase in sea level and weather disasters (13). These changes need to be monitored and controlled, as 554,000 additional malaria cases are predicted to occur between 2030 and 2049, owing to climate change. Additionally, the percentage of Africa’s population living in areas of intense malaria transmission is projected to increase from 57% to 75% as a result of climate change (14).

Asymptomatic malaria is one of three states of malaria disease, characterised by the absence of cardinal malaria symptoms, such as febrile body temperature > 37.5 °C, myalgia, and nausea; however, reduced vaccine efficacy, reduced academic performance, worsening Nodding Syndrome, and coinfections have also been reported (15–19). Approximately 85% of asymptomatic malaria cases in Ghana are caused by *P. falciparum* (20). With virtually undetectable symptoms, the infected hosts do not seek medical attention. These challenges of asymptomatic malaria give rise to a reservoir of parasitised carriers that sustain transmission in endemic regions, such as Ghana (21–23). Asymptomatic malaria infections usually occur in older children and adults in endemic regions who have developed partial immunity from continuous exposure and can persist for 18 months or more (24,25). The asymptomatic malaria pool is a significant challenge for malaria elimination (26).

Although clinical malaria cases may peak in June and July, one to two months after rainfall (2,5), asymptomatic malaria cases differ, remaining significant all year round. The prevalence of asymptomatic infections at the end of the peak malaria transmission season in school-going children is 24-27% in the costal and forest zones of Ghana (20), and higher in adults at 43-73% in the forest zone of Ghana (27,28). Outside the peak transmission season/ dry season, Abukari et al., 2019 (29) reported a 33.75% prevalence of asymptomatic parasite carriers in the coastal zone of Ghana, similar to Amoah et al., 2016 (30), conducted from February to May. However, in the northern zone, Atelu et al., 2016 (31) reported a much lower 13.9% prevalence of asymptomatic *P. falciparum* carriers during the dry season (February and March). In the wet season, a larger 82% of asymptomatic individuals in the northern zone were positive for *P. falciparum* (32).

Even with dispersed reports, it is clear that asymptomatic malaria infections are lowest during the dry season and peak at the end of the wet season in both northern and southern Ghana. This was the conclusion of Tiedje et al., 2017 (33), who investigated seasonality variation of asymptomatic malaria infection and reported a reduction in asymptomatic *P. falciparum* infections from 74.4% at the end of the wet season to 42.5% at the end of the dry season. Contrary findings of a higher prevalence in the dry season than in the wet season have also been reported by Abban et al., 2025 (34) and Ayanful-Torgby et al., 2018 (35), indicating local ecological or behavioural differences. Characteristically, most of these infections occurred in adults older than 20 years. These reports indicated an apparent lag of asymptomatic malaria infections as much as 2–5 months following the start of rainfall, and the pertinent need for specific research and strategies based on the temporal nature of the asymptomatic malaria reservoir.

Just as disease cluster and hotspot analyses advise the distribution of resources, understanding the temporal dynamics of asymptomatic malaria infections over the course of a year would advise the timing of control efforts, such as seasonal mass drug administration, and the distribution of insecticide-treated nets. Consequently, this study investigated the temporal patterns of malaria prevalence among asymptomatic parasite carriers in Simiw, Ghana, using active surveillance and meteorological data. Meteorological factors, such as rainfall, were not considered direct causes of asymptomatic malaria but represented important time points in disease seasonality.

## Materials and Methods

### Study Site

This study was conducted in Simiw, a rural town in the Komenda Edina Eguafo Abirem (KEEA) district of southern Ghana (001.33772°W and 05.16949°N). It is located in the coastal savanna ecological zone. Malaria in Simiw is seasonal, peaking in June and July (35,36). Rainfall peaks in May at the start of the transmission season (36,37).

### Study Design

This was a multi-cross-sectional study, where samples were collected from asymptomatic school-going children older than 5 years at four time points. Four community screenings were also conducted during the study. Participants were strategically recruited at time points in the peak malaria transmission season/ wet season (May 2021, August 2021), post-peak season/ wet season (October 2021), and pre-peak malaria transmission season/ dry season (February 2022), which represent the major transmission and climatic seasons in southern Ghana (Table 1). As wet or dry seasons give rise to peak and off-peak malaria transmission seasons, both types of seasons are discussed in this study.

**Table 1:** Dates of Community Screenings in Simiw.

| Month | Climate Season | Malaria Transmission Season | Dates of Community Screening |
| --- | --- | --- | --- |
| May 2021 | First Rains | Peak | May 2021 |
| August 2021 | Dry | Peak | August 2021 |
| October 2021 | Second Rains | Post-peak | October 2021 |
| February 2022 | Dry | Pre-peak | February 2022 |

Participants were considered as having asymptomatic malaria if *P. falciparum* were detected by Rapid Diagnostic Test (RDT) or Photo-induced Electron Transfer-Polymerase Chain Reaction (PET-PCR) and they lacked cardinal malaria symptoms, such as a febrile body temperature > 37.5 °C, myalgia, and nausea. Participants who had taken antimalarial drugs 2 weeks prior to sampling were excluded. Ethical clearance (073/19-20) was obtained from the Noguchi Memorial Institute for Medical Research (NMIMR) Institutional Review Board. Written consent was obtained from all the participants.

### Sample Collection

A convenience sampling approach was used, targeting school-aged children and household members. Capillary blood samples were collected by trained health workers using finger-prick procedures. Dried blood spots were prepared on filter paper for molecular analysis, and immediate diagnostic testing was performed using a Care Start™ HRP-based malaria RDT kit (Access Bio Inc, USA).

### Parasitological and Meteorological Data

DNA was extracted from dried blood spots using the Tween-Chelex DNA extraction method as previously described by Simon et al., 2020. Molecular detection of *P. falciparum* infections was accomplished by Photo-induced Electron Transfer-Polymerase Chain Reaction (PET-PCR) of the *Plasmodium falciparum* 18S ribosomal RNA gene (*Pf*s18s) gene, as described by Matamoros et al., 2023 (38). Meteorological data for the KEEA district from March 2021 to February 2022 was obtained from the Ghana Meteorological Agency. Meteorological data included atmospheric temperature, precipitation, and humidity.

### Statistical Analysis

Descriptive statistics were calculated. Analysis of Variance (ANOVA) was used to test the difference between the median ages of participants who were *P. falciparum* positive and *P. falciparum* negative, controlling for wet/ dry season. Logistic regression was used to determine the odds of a *P. falciparum* infection given age, controlling for season.

For analysis, meteorological data was aggregated monthly by total monthly precipitation, mean daily relative humidity, mean daily average temperature, mean daily maximum temperature, and mean daily minimum temperature. A sequence chart of meteorological and asymptomatic infections during the year was used to determine visual trends of asymptomatic malaria cases. Spearman’s rank correlation test was used to assess the pairwise relationships between meteorological variables and counts of asymptomatic malaria cases. Poisson Generalised Linear Modelling with splines was used to analyse the nonlinear relationship between asymptomatic malaria cases as the outcome and seasonal trend and meteorological variables as predictors. In disease epidemiology, there are three types of trends in disease occurrence. These include short-term fluctuations (i.e., epidemics), periodic fluctuations (i.e., seasonal trends for most communicable diseases and cyclic trends over a few years), and secular/ temporal trends (i.e., over decades) (39,40). In the present study, the seasonal trend of asymptomatic infections was studied. It encompasses factors that vary or have a pattern over time in the *Plasmodium*-human ecosystem.

Distributed Lag Nonlinear Modelling (DLNM) was used to analyse the lagged nonlinear influence of meteorological variables as predictors of asymptomatic malaria cases. For model simplicity, both the degrees of freedom and maximum lag were set to two (2). The effects of variables, while holding the other variables constant at their means, were isolated in all models. These were expressed as exponentiated coefficients (exp(β)), representing multiplicative changes in the outcome. For example, exp(β) = 1.20 for rainfall indicates a 20% increase (or 1.2x) in the expected number of cases per one-unit increase in rainfall (mm), the predictor.

Asymptomatic cases, as determined by PET-PCR, were used for ANOVA, logistic regression, and all models. Statistical analysis was performed using R version 4.4.2 and the α-value was set at 0.05.

## Results

### Demographics of Study Participants

At the end of the study period, 846 asymptomatic participants had been recruited; approximately 200 school-going children and community members at every time point. The ratio of males to females was approximately 1:3. The median age of the participants was 36 years. The average body temperature and haemoglobin level of the asymptomatic participants were 36.5 °C and 12.1 g/dL respectively. Asymptomatic *P. falciparum* infections ranged from 6.2% to 14% by RDT and 13.4% to 32% by PET PCR (Table 2).

**Table 2:** A Table of Participant Demographics.

| Characteristic | Sampling Time |  |  |  |  |
| --- | --- | --- | --- | --- | --- |
|  | May 2021 | August 2021 | October 2021 | February 2022 | Total |
| No. of participants | 200 | 209 | 210 | 226 | 845 |
| Male/ Female | 59/ 139 | 61/ 148 | 45/ 165 | 57/ 169 | 222/621 |
| Median age (IQR) | 30.5 (13.75-46.25) | 30 (15-52.5) | 41 (26-55) | 40 (30-56) | 36 (17-52) |
| Median Body Temperature (°C) (IQR) | 36.5 (36.4-36.6) | 36.2 (36.1-36.3) | 36.7 (36.5-37) | 36.6 (36.5-36.9) | 36.5 (36.3-36.7) |
| Median Haemoglobin Level (g/dL) (IQR) | 12.1 (10.9-13.6) | 12 (11.2-13.5) | 12 (11-13) | 12.1 (11-13.4) | 12.1 (11-13.3) |
| RDT *P. falciparum* Positive/ Total | 17/ 200 (8.5%) | 13/ 209 (6.2%) | 29/ 210 (14%) | 14/ 226 (6.2%) | 73/ 845 (8.0%) |
| PET-PCR *P. falciparum* Positive/ Total | 44/ 156 (22.0%) | 28/ 209 (13.4%) | 51/ 210 (24.3%) | 72/ 225 (32.0%) | 195/ 800 (24.4%) |

No. = Number; IQR = Interquartile Range

There was a statistically significant difference between the median ages of participants who were *P. falciparum* positive (14 years (IQR = 9.5-32.5 years)) and *P. falciparum* negative (40.5 years (IQR = 26-56 years)) (p < 0.0001) (Fig 1). Consequently, every one-year increase in age was associated with a 6% reduction in the odds of *P. falciparum* infection (OR = 0.944, p < 0.0001).

**Fig 1:**
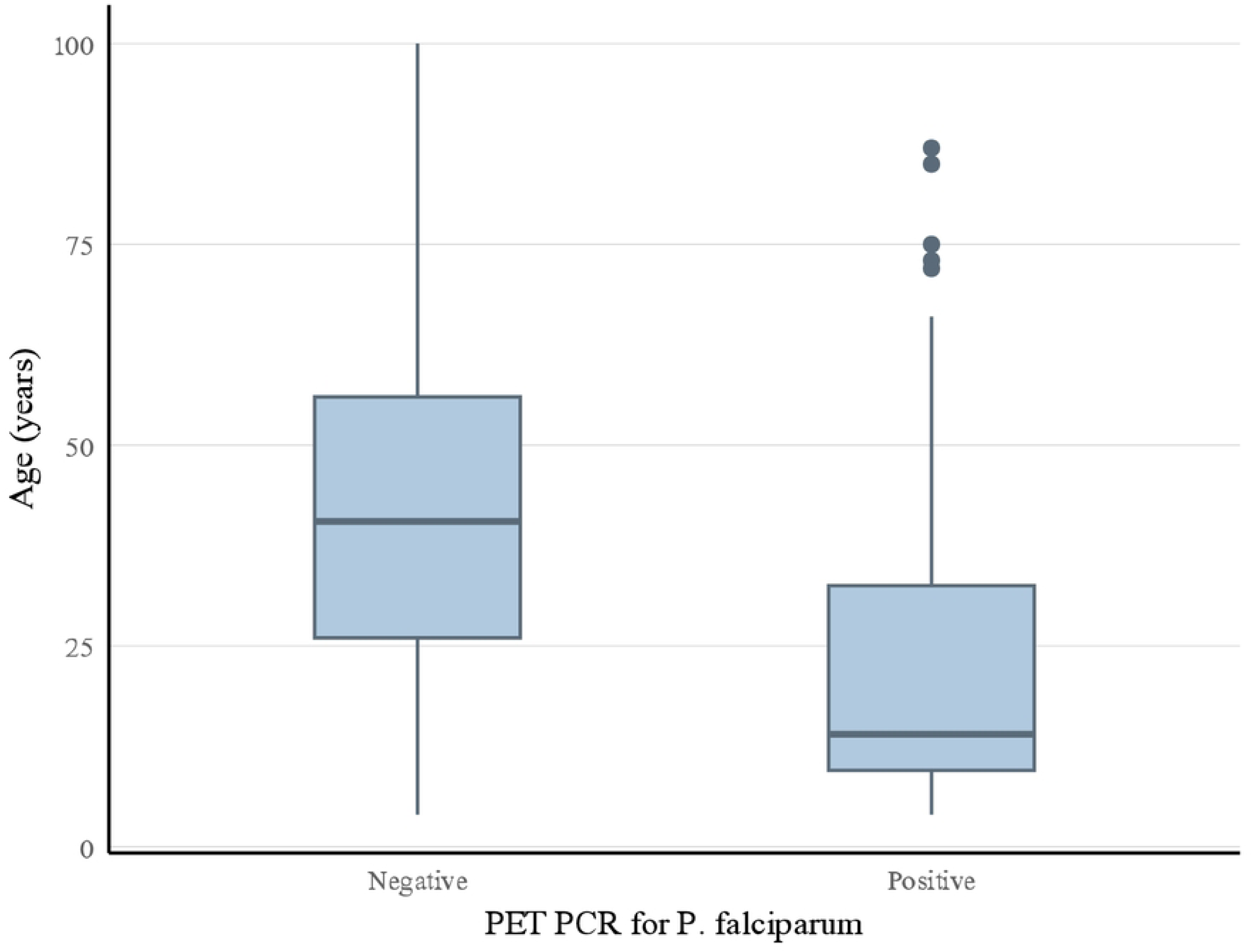
A box plot of age distribution against asymptomatic *P. falciparum* infection.

### Meteorological Trends and Temporal Changes of Asymptomatic *P. falciparum* Infections

The highest prevalence of asymptomatic infections determined by PCR (32.0%) and the lowest prevalence of asymptomatic infections determined by RDT (6.2%) occurred during the dry season in February 2022. The lowest prevalence of asymptomatic infections, as determined by both PCR (13.4%) and RDT (6.2%), occurred in August 2021, during the first rainy season (Table 2 and Fig 2). Interesting, the lowest prevalence of infections determined by RDT occurred in both August 2021 and February 2022 with the same value, 6.2%. The second highest prevalence of asymptomatic infections determined by PCR (24.3%) occurred in October 2021, followed closely by May 2021 (22.0%). Oddly, in August, between May and October, Simiw saw the lowest dip in asymptomatic infections by both PCR and RDT. (Fig 2).

**Fig 2:**
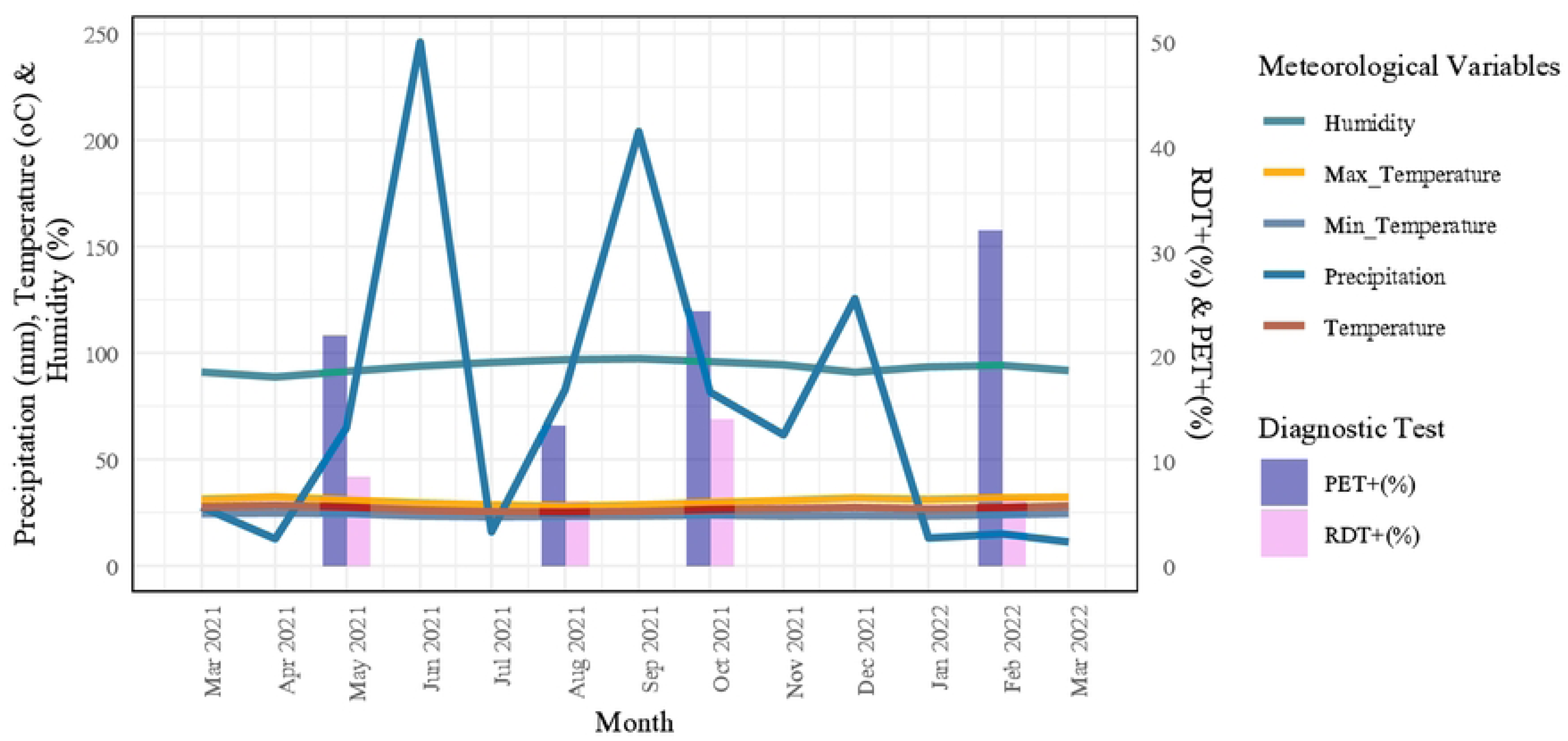
A sequence chart of meteorological trends and asymptomatic malaria counts in Simiw. The months of the year are represented on the x-axis. The total monthly precipitation (light blue), temperature (average, minimum, and maximum; red, blue, and yellow, respectively), and humidity are represented by line graphs and read on the left axis. Percentage prevalence of *P. falciparum* infections by RDT (pink) and PET-PCR (lavender) are represented by bar graphs and read on the right axis. Diagnostic tests were performed in May 2021, August 2021, October 2021, and February 2022 only.

Visually, the trend of asymptomatic *P. falciparum* prevalence, as detected by RDT, appeared to follow the total monthly precipitation with a lag of one month. However, the prevalence of asymptomatic *P. falciparum* infections, as detected by PET-PCR, seems to follow the total monthly precipitation with a lag of 3 months (Fig 2).

Changes in atmospheric temperature and humidity occurred within a narrow range, and patterns with asymptomatic *P. falciparum* infections were not observed. During the study period, the average, minimum, and maximum atmospheric temperatures remained fairly constant at 26.8°C (26.2°C-27.4°C), 23.5°C (23.3°C–24.2°C) and 30.9°C (29.4°C–31.5°C), respectively (interquartile ranges are in parentheses). The total precipitation per month seemed to fluctuate with the two wet seasons, with an annual average of 61.5 mm (15.1 mm–82.1 mm). The highest amount of precipitation of 246.1 mm was recorded at the end of the first rainy season, June 2021, and the lowest amount of precipitation of 11.2 mm was recorded at the end of the dry season, March 2022. The average humidity was 90% (90%-90%), with a maximum humidity of 100% occurring in July, August, and September 2021 (Fig 2).

Statistically, the nonlinear relationship of time with asymptomatic malaria counts was statistically significant (exp(β) = 190.377; p = 0.0109) (Fig 3), rising in June 2021 of the wet season and peaking at the start of October 2021, after the first rainy season and at the onset of the second rainy season. The modelled time-trend peak of asymptomatic cases in October 2021 differed from the field-observed peak in February 2021 because the model estimated the expected counts over time after smoothing (natural spline) and holding covariates at their means. Smoothing and covariate adjustment shifted the fitted peak away from the raw data.

**Fig 3:**
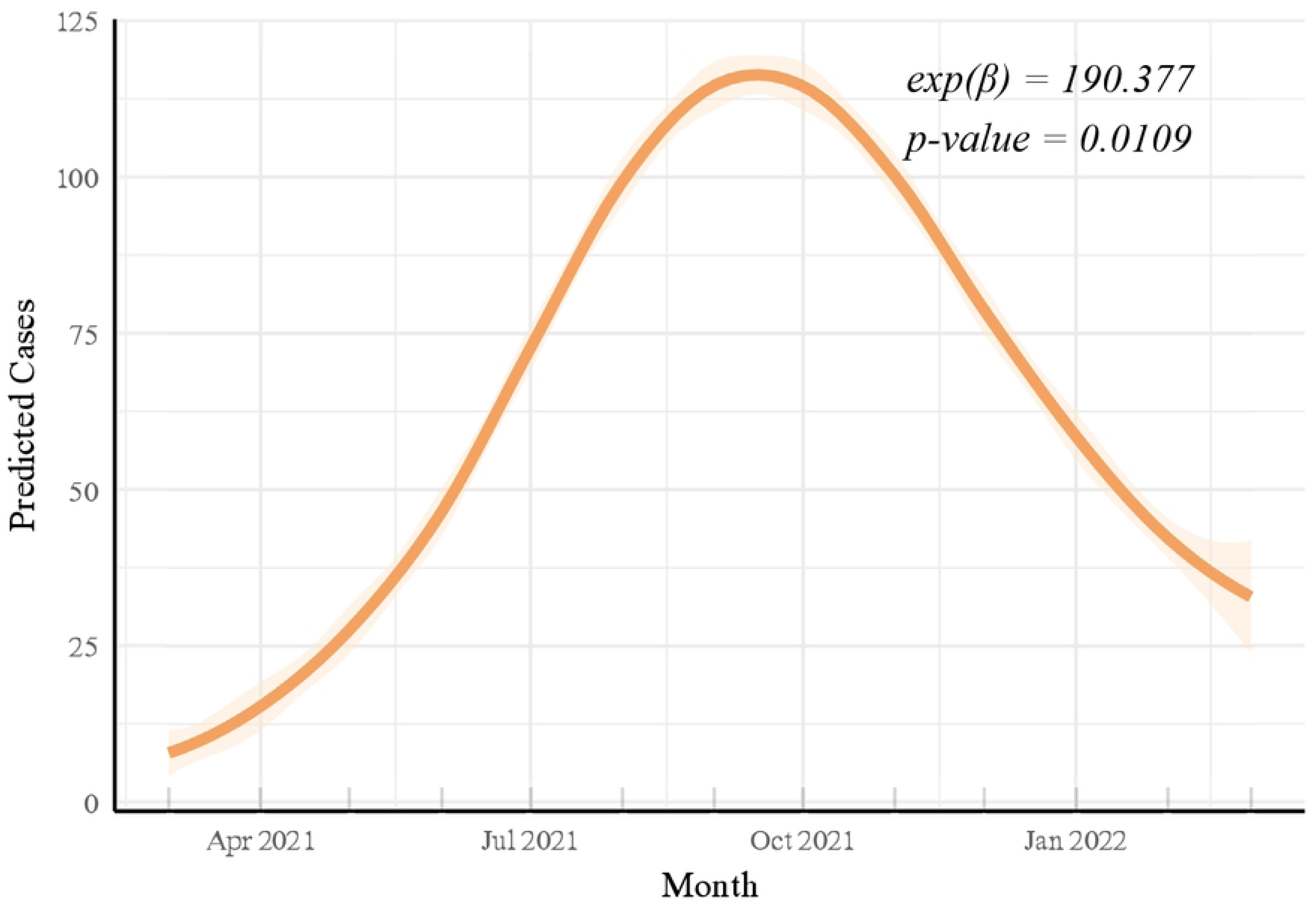
A line plot of the nonlinear relationship between time and asymptomatic *P. falciparum* infections. This line plot shows the modelled association between time, represented by months of the calendar year, and predicted counts of asymptomatic malaria cases. It was produced by Poisson Generalised Linear Modelling with a natural spline applied to time (degrees of freedom is 2), and other covariates held at their means. It revealed a bell-shaped pattern, rising in June and peaking around October. The shaded band represents the 95% confidence interval.

There was a statistically significant nonlinear relationship between humidity (exp(β) = 0.734; p = 0.0161) (Fig 4), maximum atmospheric temperature (exp(β) = 1.301, p = 0.000109) (Fig 5) and precipitation (exp(β) = 0.99; p = 0.0000571) (Fig 6). The lowest number of predicted asymptomatic malaria cases occurred at 250 mm precipitation and 97.5% atmospheric humidity, while the highest count of asymptomatic malaria cases occurred at a maximum temperature of 32°C. There was no statistically significant linear correlation between any meteorological variable and the number of asymptomatic infection cases (p > 0.76).

**Fig 4:**
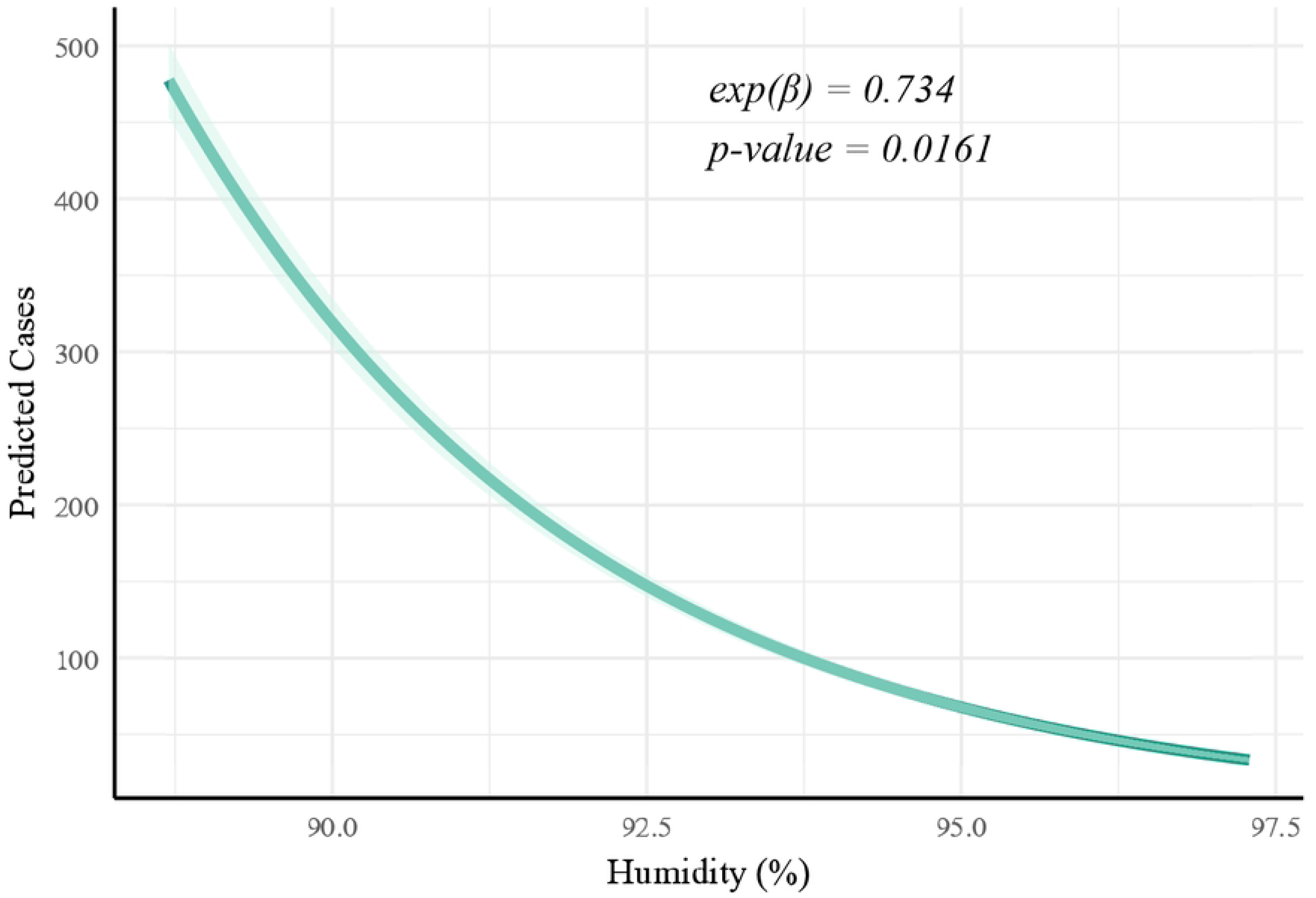
A line plot of the nonlinear relationship between humidity and asymptomatic *P. falciparum* infections. This line plot shows the modelled association between humidity and predicted counts of asymptomatic malaria cases. It was produced by Poisson Generalised Linear Modelling with a cross-basis natural spline applied to humidity, and other covariates held at their means. It revealed a sloped curve that decreases sharply from approximately 500 predicted cases at 85% and reaches near 0 at 97.5%. The shaded band represents the 95% confidence interval.

**Fig 5:**
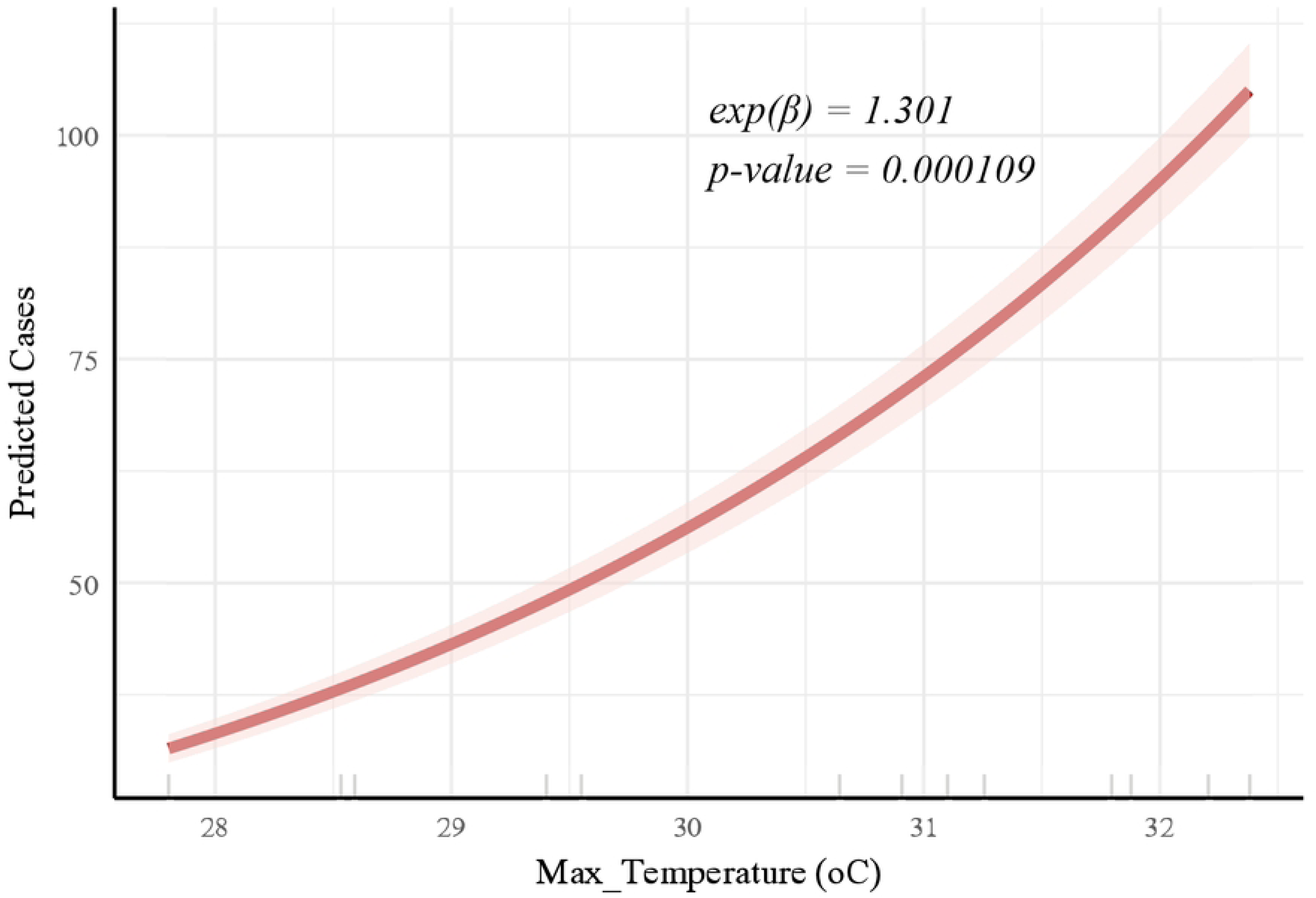
A line plot of nonlinear relationship between maximum atmospheric temperature and asymptomatic *P. falciparum* infections. This line plot shows the modelled association between maximum atmospheric temperature and predicted counts of asymptomatic malaria cases. It was produced by Poisson Generalised Linear Modelling with a cross-basis natural spline applied to the maximum atmospheric temperature, and other covariates held at their means. It revealed a sloped curve that rises sharply from near 0 at 28°C and reaches approximately 100 predicted cases at 32.5°C. The shaded band represents the 95% confidence interval.

**Fig 6:**
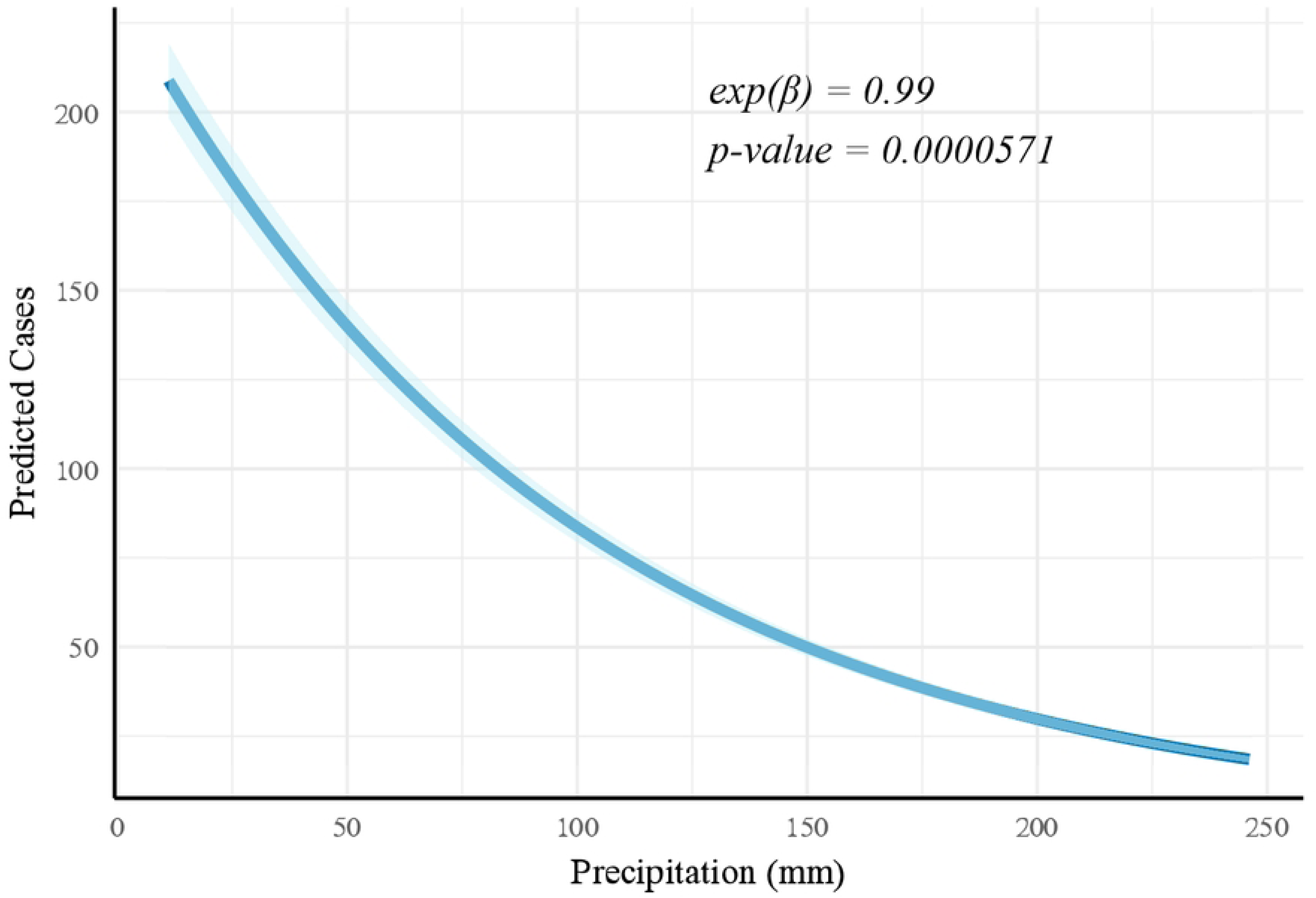
A line plot of the nonlinear relationship between precipitation and asymptomatic *P. falciparum* infections. This line plot shows the modelled association between precipitation and predicted counts of asymptomatic malaria cases. It was produced by Poisson Generalised Linear Modelling with a cross-basis natural spline applied to precipitation, and other covariates held at their means. It reveals a sloped curve that decreases gradually from approximately 200 predicted cases at 0 mm and reaches near 0 predicted cases at 250 mm. The shaded band represents the 95% confidence interval.

When meteorological variables were lagged, only precipitation was statistically significant at lags of one (1) month (exp(β) = 0.252; p = 0.0306) and two months (2) (exp(β) = 3.202; p = 0.004) (Fig 7). Humidity and atmospheric temperature (average, maximum, and minimum) were not statistically significant at lags of one (1) or two (2) months (p > 0.05).

**Fig 7:**
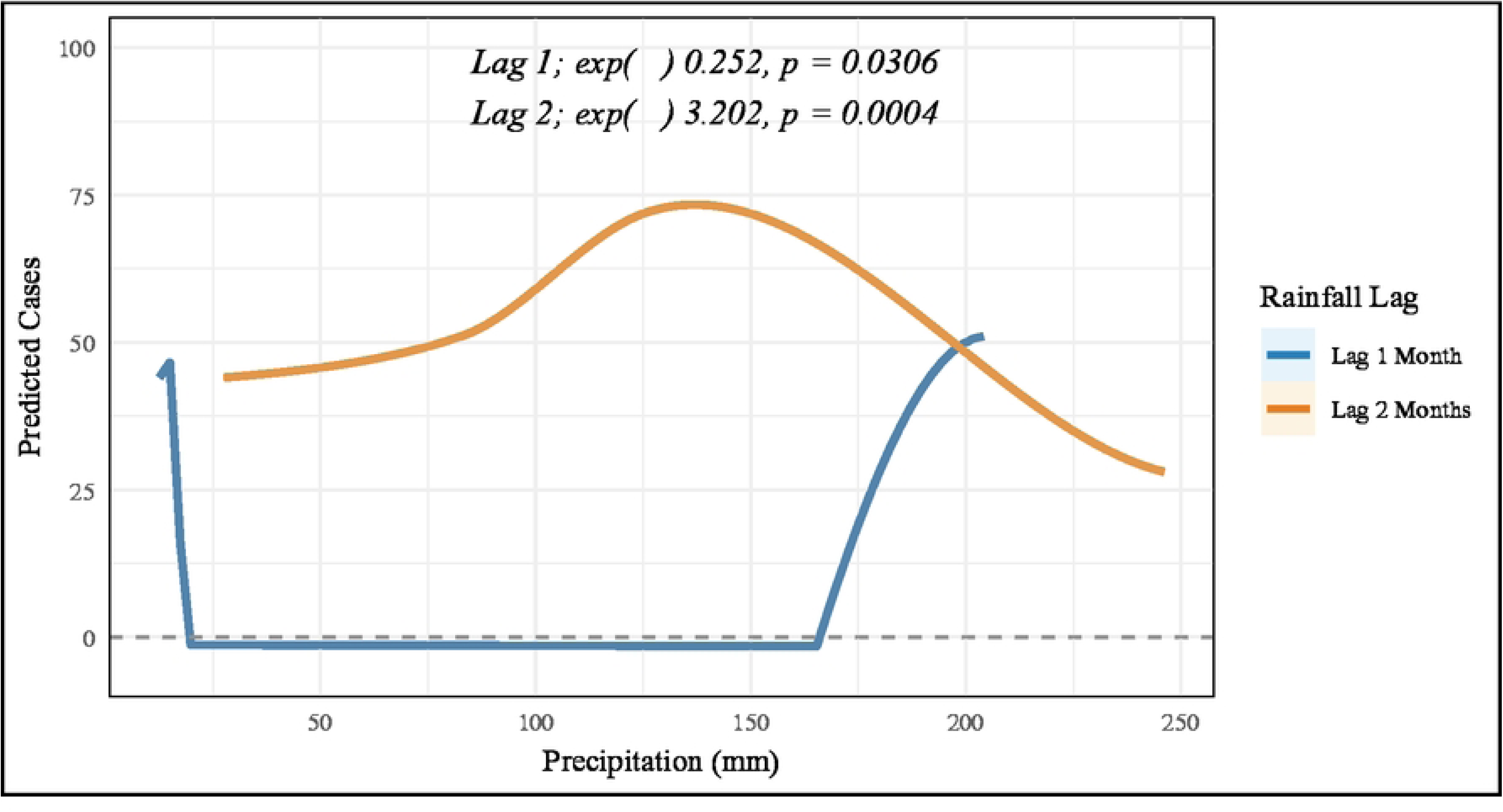
A line plot of the lagged relationship between precipitation and asymptomatic *P. falciparum* infections. This line plot shows the modelled association between precipitation at lags 1 and 2 months and the predicted counts of asymptomatic malaria cases. It was produced by Distributed Lag Nonlinear Modelling with a cross-basis natural spline function for precipitation. Predictions were generated using crosspred with other covariates held at their means. At a lag of 1 month (blue line), the predicted cases instantly drop below 0 at approximately 0 mm, and only start to rise from 0 predicted cases at approximately 150 mm to 50 predicted cases at 200 mm. At a lag of 2 months (orange line), the predicted cases were steady at 50 cases from 50 mm, peaking at approximately 150 mm with 75 predicted cases, and dipping to 25 predicted cases at 250 mm.

## Discussion

The National Malaria Elimination Strategic Plan (NMESP) 2024-2028 of Ghana, based on the World Health Organisation (WHO) Global Technical Strategy (GTS) for Malaria 2016–2030, aims to eliminate malaria in Ghana (41,42). Accomplishing this goal will require an understanding of the temporal dynamics of malaria in the asymptomatic pool of carriers that sustains year-round transmission. Malaria transmission is seasonal, mostly because vector populations depend on climatic factors that cycle over time. As such, the incidence of clinical malaria cases is seasonal. However, this is not the case for asymptomatic malaria infections, which do not arise directly from vector inoculations but persist all year-round following reduced parasite virulence and immunological responses (host-parasite equilibration). In the present study, statistical modelling was used to delineate the temporal properties of asymptomatic malaria prevalence in the endemic community of Simiw, Ghana, over one year. The influence of climatic factors was also examined, owing to their seasonal properties.

In this study, the highest prevalence of asymptomatic infections, as determined by PCR (32.0%), occurred in February during the dry season. Ironically, this month recorded the lowest prevalence of asymptomatic infections, as determined by RDT. This indicates an increase in low-density infections and a decrease in high-density infections during the dry season, consistent with the decreased parasite virulence during the low malaria transmission season (26). October, the start of the second wet season, saw the second highest prevalence of asymptomatic infections determined by PCR (24.3%), followed closely by May (22.0%), the first rainy season. This suggests that some symptomatic infections, accumulated from the vector rise during the first rainy season, may have transitioned into low-density asymptomatic infections after 6 months. Interestingly, Coulibaly et al., 2017 (43) found no evidence that asymptomatic malaria infections lagged behind clinical cases but indicated that they may lead/ precede them, seeding clinical infections at the start of the rainy season. These findings indicate the persistence of asymptomatic infections throughout the year, in both high and low transmission seasons. In low transmission seasons, most infections become low parasite density, possibly to reduce survival expenditure, persisting for months until the vector rises in the wet season (44,45). These low-density infections would then seed new infections once the rains return in the wet season, leading to a rise in clinical cases (11,46).

Remarkably, the prevalence of asymptomatic cases in Simiw was higher in the dry season than in the wet season, when clinical cases gain their peak (11). This represents a shift in the peaks of clinical malaria in the first wet season (11,46) and aclinical malaria cases in the dry season. A higher prevalence of asymptomatic *falciparum* infections in the dry season than in the wet season has also been reported in Ghana by Abban et al., 2025 (34) and Ayanful-Torgby et al., 2018 (35). Additionally, the ratios of asymptomatic cases to clinical cases in the dry and wet seasons have been reported to be 5:1 and 0.5:1, respectively (43), further supporting the findings of the present study. The higher proportion of asymptomatic infections in the dry season may be an adaptation of the parasite for survival during low malaria transmission (44,47). Throughout the year, most asymptomatic *P. falciparum* infections in Simiw were recorded in older children (median age: 14 years (IQR = 9.5-32.5 years)), consistent with the findings of Kołodziej et al., 2024 (24). This may be because of partial immunity in older children, as well as the immunological experience and healthier behaviours among adults, such as consistent bed net use, indoor living, and access to healthcare.

Having reported the overall pattern of asymptomatic malaria infections in Simiw, we examined the seasonal trend of asymptomatic malaria, where time and meteorological variables were isolated and held at their means. The nonlinear relationship between time and asymptomatic malaria counts was bell-shaped, peaking around October 2021, after the first rainy season and at the onset of the second rainy season. The modelled time-trend peak of asymptomatic cases in October 2021 differed from the field-observed peak in February 2021 because the model estimated the expected counts over time after smoothing (natural spline) and holding covariates at their means. Smoothing and covariate adjustment shifted the fitted peak away from the raw data. Even so, the model predicted the highest asymptomatic malaria infections more than five months after the first rains, similar to the field observation in Simiw. While clinical malaria cases lag behind the first rainy season by more than one month accounting for vector breeding, infective bites and the development of the asexual parasite densities in the host (48–54), the longer lag of asymptomatic infections likely accounts for same factors and an additional host–parasite equilibration during the infection. Host–parasite equilibration, known as malaria premunition, may be driven by reduced parasite virulence, partial immune control, and incomplete treatment from repeated parasite exposures that favour low-cost persistence in the asymptomatic host.

The bimodal climate of Simiw is favourable for malaria transmission, as *Anopheles* mosquitoes thrive at 18-32°C and > 60% humidity (55,56). This may not be the case for asymptomatic malaria, which is not directly transmitted by the vector but develops over time following reduced parasite virulence and host immunological response. That said, there appears to be a minor meteorological influence on asymptomatic infections, probably within a composite epidemiological system. Several studies have reported varying effects of meteorological variables on the malaria burden. Generally, features of the wet season, such as rising rainfall and humidity, increase malaria transmission and clinical cases in most regions (43,57–61). However, in the present study, features of a dry season, such as decreasing rainfall and rising maximum atmospheric temperature, increased cases of asymptomatic malaria infections. According to the models, the highest counts of asymptomatic malaria cases occur at a maximum daily temperature of 32°C, daily relative humidity of 91% and monthly precipitation of 20 mm, similar to records in February 2022 in the dry season. Usually, high temperatures are fatal to mosquito vectors (62,63). However, this may not limit asymptomatic malaria, which persists with or without the vector. Humidity decreased asymptomatic malaria cases in Simiw, just as it does for clinical malaria (59).

Meteorological variables have different lagged effects on malaria epidemiology. As with clinical malaria cases that lag rainfall by zero to six (0–6) months because of the life cycle of the mosquito (48–54), asymptomatic malaria cases lagged precipitation by one to two (1–2) months in Simiw, similar to Das et al., 2015 (64). Despite being in the lag range of clinical malaria cases (0-6 months lag), precipitation is likely a proxy for more relevant temporal factors. The peak of this effect occurs at 150 mm, far below the normal precipitation of 800 mm in the region (9); essentially a dry season optimal for asymptomatic parasite persistence. Based on these findings, mass drug administration in this community can be implemented two months after ∼150 mm rainfall in the dry season (65).

The seasonal influence of time on asymptomatic malaria infections in Simiw was significantly higher than meteorological variables. This is expected, as vectors do not specifically transmit asymptomatic malaria. Unlike clinical malaria, which is strongly influenced by meteorological factors, the stronger effect of time indicates that host and parasite factors shape annual asymptomatic malaria patterns. These likely include agricultural labour patterns, indoor residual spraying schedules, seasonal migration, health-seeking behaviour, seasonal immune modulation, and malaria control schedules. For example, human nocturnal activity has been linked to asymptomatic *P. falciparum* carriage in epidemiological studies in Ghana (56,66,67). It is no coincidence that *Anopheles* mosquitoes, being photophobic, exhibit night-biting behaviour (68–70). In fact, many host behaviours shift throughout the year because of festivals, occupation, and seasonal malaria control strategies, among others. Additionally, the circannual rhythms of host immunity could also explain trends in asymptomatic malaria (71,72). Seasonal modulation has been reported as immunological gene expression varies within and between seasons (73,74). This modulation has been shown to be an independent driver of the seasonality of respiratory infections (73). Furthermore, it has been theorised that the parasite in an infected host may sense host physiological or behavioural changes during the wet season (44).

## Conclusion

In this study, asymptomatic malaria cases were detected all year round, peaking in the dry season. Interestingly, low-density asymptomatic infections were higher than high-density infections in the dry season. Such a protracted transmission profile implies that parasites persist during intervening gaps of transmission seasons, maintaining an asymptomatic reservoir in Simiw. As expected, the seasonal influence of time on asymptomatic malaria was stronger than climatic factors. Asymptomatic infections were positively associated with the maximum atmospheric temperature and negatively associated with precipitation and humidity, as expected in the dry weather conditions of low malaria transmission. A precipitation of 150 mm was also associated with asymptomatic malaria cases at lags of 1 and 2 months. This finding is relevant, as timely and accurate information about the onset of asymptomatic malaria surges in endemic regions is essential for effective malaria control activities. It may also provide early warning methods of surges of asymptomatic malaria cases in endemic regions.

Malaria temporal epidemiology is understudied. We found only two studies that focused on asymptomatic malaria infections (43,64). Therefore, we recommend more studies on this subject; specifically on inter-year variability, secular trend, and geographical differences (52). A limitation of this study is the sparsity of asymptomatic *P. falciparum* infection data.

## Declaration

### Authors’ Contributions

Conceptualization: LEA, AP, RT, CKMAA,

DA Data Curation: FKA, EKO, DA, CKMAA

Formal Analysis: CKMAA

Funding Acquisition: LEA, AP, RT

Investigation: FKA, EKO, DA

Methodology: LEA, AP, RT, CKMAA, DA

Project Administration: LEA

Resources: LEA, AP, RT

Software: CKMAA

Supervision: LEA, AP, RT

Validation: FKA, DA

Visualization: CKMAA

Writing – Original Draft Preparation: CKMAA

Writing – Review & Editing: CKMAA, LEA, FKA, EKO, AP, RT, DA

## Data Availability

For ethical reasons and protection of participant data, contact the corresponding author to make available data when needed.

## Acknowledgment

Not Applicable

